# Cerebral Microbleeds and the Effect of Early Antihypertensive Treatment in Patients with Acute Ischemic Stroke: A Secondary Analysis of the CATIS-2 Trial

**DOI:** 10.1101/2025.08.13.25333633

**Authors:** Zilin Zhao, Xuewei Xie, Yuesong Pan, Mengxing Wang, Yufei Wei, Ximing Nie, Aili Wang, Dacheng Liu, Wanying Duan, Xin Liu, Zhe Zhang, Jingyi Liu, Lina Zheng, Minghua Wang, Yong Jiang, Jing Jing, Xia Meng, Katherine Obst, Chung-Shiuan Chen, Hao Li, David Wang, Yilong Wang, Yonghong Zhang, Jiang He, Yongjun Wang, Liping Liu, the CATIS-2 Investigators

## Abstract

**Background and Objectives:** To explore whether the presence, burden or distribution of cerebral microbleeds (CMBs) modifies the effect of early versus delayed antihypertensive treatment on clinical outcomes in acute ischemic stroke (AIS) patients.

**Methods:** A secondary analysis of the China Antihypertensive Trial in Acute Ischemic Stroke II (CATIS-2) trial was performed. Participants with baseline cerebral magnetic resonance imaging (MRI) data for CMB evaluation were included. The primary outcome was functional dependency or death at 90 days, defined as a modified Rankin Scale (mRS) score ≥3.

**Results:** Among 815 eligible participants (mean age 62.4 years; 34.5% female), 396 (48.6%) had at least one CMB. The presence of strictly deep CMBs was associated with an increased risk of functional dependency or death at 90 days compared to the absence of CMBs (15.3% vs 7.9%; adjusted odds ratio [aOR] 1.92, 95% CI 1.08-3.40, *P*=0.03). Early versus delayed antihypertensive treatment significantly increased the risk of functional dependency or death in patients with CMBs (17.1% vs 8.7%; aOR 2.17, 95% CI 1.16-4.07; *P*=0.02). This effect was particularly pronounced among patients with moderate-to-severe CMBs (17.5% vs 6.6%; aOR, 2.85; 95% CI, 1.06–7.67; *P*=0.04) and strictly deep CMBs (20.8% vs 9.0%; aOR, 2.76; 95% CI, 1.15–6.64; *P*=0.02). However, no statistically significant interaction was observed between CMBs and treatment assignment for the primary outcome (*p*_interaction_>0.05).

**Conclusions:** This secondary analysis of CATIS-2 indicates that the presence of CMBs, particularly those predominantly located in deep regions or with a greater burden, may increase the risk of adverse clinical outcomes following early antihypertensive treatment in AIS patients. These findings highlight the imperative for further research on individualized blood pressure (BP) management strategies in AIS patients with CMBs.

**Registration:** URL: https://www.clinicaltrials.gov; Unique identifier: NCT03479554.

## Introduction

Elevated blood pressure (BP) is commonly observed during the acute phase of ischemic stroke and is associated with poor functional prognosis and an increased risk of ischemic stroke recurrence.^1–3^ Despite extensive research, the optimal timing for initiating antihypertensive treatment in patients with acute ischemic stroke (AIS) remains controversial. Evidence to date suggests that early BP reduction following AIS may exert neutral or even detrimental effects on clinical outcomes,^4–6^ underscoring the need for tailored rather than generalized BP management strategies.

Advancements in neuroimaging have facilitated the identification of specific markers, such as cerebral microbleeds (CMBs), which have garnered attention due to their potential influence on the clinical outcomes of ischemic stroke. CMBs are small, hypointense lesions visible on hemorrhage-sensitive magnetic resonance imaging (MRI) sequences, and are recognized as radiological markers of cerebral small vessel disease (CSVD).^7^ CMBs are highly prevalent in ischemic stroke patients,^8,9^ are strongly associated with hypertension,^10,11^ and are predictive of adverse functional outcomes.^8,12–14^ Moreover, there is evidence suggesting that patients with CMBs may exhibit impaired cerebral blood flow autoregulation,^15,16^ raising concerns that they may be particularly vulnerable to the adverse effects of early BP-lowering interventions.

To date, no studies have examined whether the presence, burden, or distribution of CMBs modifies the optimal timing for BP reduction in AIS. The China Antihypertensive Trial in Acute Ischemic Stroke-2 (CATIS-2), a multicenter randomized controlled trial, demonstrated that early initiation of antihypertensive treatment (within 24–48 hours) did not reduce the odds of dependency or death at 90 days compared with delayed treatment (initiated on day 8).^17^ It remains plausible that neuroimaging markers of CSVD, such as CMBs, may help identify subgroups of patients who respond differently to early BP reduction strategies. To address this hypothesis, we conducted a secondary analysis of CATIS-2 to evaluate the association between CMBs and functional prognosis and to explore whether CMBs modify the response to early versus delayed antihypertensive treatment in AIS patients.

## Methods

### Design and Participants

The rationale, design, participant characteristics, and main results of the CATIS-2 trial have been previously reported.^17,18^ Briefly, CATIS-2 was a multicenter, randomized, open label, blinded-endpoints clinical trial conducted in 106 hospitals across China from June 2018 through July 2022. It compared the impact of early antihypertensive treatment versus delayed treatment on reducing dependency or death in patients with AIS (trial protocol and statistical analysis plan in Supplemental Material.). Eligible participants were patients aged 40 years or older with AIS (symptom onset within 24-48 hours) and elevated systolic BP (SBP) between 140 and 220 mm Hg. Patients with atrial fibrillation, severe stroke (National Institutes of Health Stroke Scale (NIHSS) score ≥21), and those treated with intravenous thrombolytic treatment or endovascular thrombectomy were excluded.

Our study included CATIS-2 participants with evaluable cerebral microbleeds (CMBs), defined as those whose baseline MRI included a readable axial susceptibility-weighted imaging (SWI) or T2*-weighted gradient echo (GRE) sequence allowing for CMB detection. Thirty CATIS-2 participating centers had complete imaging data meeting these criteria and thus were included in the current substudy.

### Standard Protocol Approvals, Registrations, and Patient Consents

The trial was approved by the ethics committees of Beijing Tiantan Hospital, all participating institutes in China, and the Institutional Review Board of Tulane University in the US. Informed consent was signed by patients or their representatives before enrollment. The CATIS-2 trial was registered at ClinicalTrials.gov (Unique identifier: NCT03479554).

### Intervention

Patients were randomly assigned (1:1) to receive antihypertensive treatment immediately after randomization (aimed at reducing SBP by 10%-20% within the first 24 h and a mean BP <140/90 mm Hg within seven days) or to discontinue antihypertensive medications for seven days, followed by treatment initiation on day 8 (aimed at achieving mean BP <140/90 mm Hg). A randomization sequence was generated centrally using a web-based system managed by the Study and Data Coordinating Center at Beijing Tiantan Hospital.

### Data Collection

Demographic characteristics and medical history were collected at enrollment. Stroke severity was evaluated using the NIHSS score by trained neurologists at admission, 14-day or hospital discharge, and 90-day follow-up visits. BP measurements were conducted by trained nurses using an automated device (Omron HBP-1300 Professional BP Monitor) according to a standard protocol recommended by the American Heart Association.^19^ Three BP measurements were obtained at baseline; every 3 hours after randomization for the first 24 hours; every 8 hours from day 2 until day 14 or hospital discharge; and at 21-day and 90-day follow-up visits.

### Imaging Acquisition and Analysis

Brain MRI scans of enrolled patients were performed according to clinical judgment at the individual centers, with these decisions made independently of treatment assignment. And imaging data were centrally collected in Digital Imaging and Communications in Medicine (DICOM) format from individual centers. Centralized interpretation of the imaging data was independently performed by two experienced neuroradiologists blinded to clinical data. A third neuroradiologist was involved for additional assessment in cases of disagreement.

CMBs were defined as small (2-10 mm in diameter) round or oval hypointense lesion on SWI or GRE sequences, with associated halos that are not visible on T1- or T2-weighted images.^7^ CMBs were classified as present or absent, and if present, further categorized as strictly deep (deep and/or brainstem, cerebellar CMBs), strictly lobar or mixed (concurrent deep and lobar CMBs). Severity (CMB burden) was classified as absent (0 CMBs), mild (1-2 CMBs), or moderate to severe (≥3 CMBs).^20^

Lacunes, white matter hyperintensities (WMHs) and enlarged perivascular spaces (EPVSs) were evaluated using consensus criteria.^21^ WMHs were graded using the Fazekas scale, with the scores for periventricular and deep WMHs combined to obtain the total Fazekas score. Moderate-to-severe WMH burden was defined as a total Fazekas score between 3 and 6. EPVSs in the basal ganglia and centrum semiovale were graded on a validated 4-point scale in T2-weighted sequences, with moderate-to-severe burden defined as >10 spaces in either location.

Intracranial atherosclerotic stenosis (ICAS) was assessed based on the Warfarin-Aspirin Symptomatic Intracranial Vascular Disease (WASID) trial criteria, with the presence of ICAS defined as ≥50% stenosis in any of the following arteries on magnetic resonance angiography (MRA): intracranial internal carotid artery, middle cerebral artery (M1/M2 segment), anterior cerebral artery, posterior cerebral artery, intracranial vertebral artery, or basilar artery.

### Outcomes

The primary outcome of the current study was the composite outcome of death or functional dependency (modified Rankin Scale [mRS] score ≥3) at 90 days after randomization. Secondary outcomes included ordinal mRS scores, recurrent stroke and major vascular disease events (ie, vascular deaths, non-fatal stroke, non-fatal myocardial infarction, coronary revascularization, and hospitalized angina and heart failure) at 90 days. The outcomes were ascertained through follow-up visits by trained neurologists who were blinded to the treatment assignments.

### Statistical Analysis

Intent-to-treat analyses were used to assess outcomes in patients with available clinical endpoint information. Continuous variables are presented as mean with standard deviation (SD) or median with interquartile range (IQR), and categorical variables are presented as frequency and percentage. Baseline characteristics between the early and delayed treatment groups were compared using a *t* test or Kruskal-Wallis test for continuous variables and a χ^2^ test or Fisher’s exact test for categorical variables. Logistic regression analysis was used to estimate odds ratios (ORs) and 95% CIs associated with early treatment compared with delayed treatment. Ordinal logistic regression was used to estimate the effect of early versus delayed treatment on the full range of the mRS. In addition, the heterogeneity of the treatment effect on clinical outcomes based on CMB presence, burden and topography was assessed by adding an interaction term (CMBs × early antihypertensive treatment) in the logistic regression models. Linear mixed-effects regression analysis was used to test the group differences in mean BP changes between the early and delayed treatment groups with a significance level of 0.004 (0.05/14 tests). Statistical significance was defined by *p* < 0.05. All analyses were two-sided and performed using SAS software version 9.4 (SAS Institute, Inc., Cary, NC).

### Data Availability

The data that support the results of this study are accessible from the corresponding author for reasonable research purposes.

## Results

### Patient Demographics and Baseline Characteristics

A total of 815 patients who underwent MRI scans containing SWI/GRE sequence were included in the current subgroup analysis (Figure S1 in Supplement 3). The mean (SD) age of the included patients was 62.4 (9.7) years, and 535 (65.6%) were male. The baseline characteristics for the patients enrolled and excluded in this analysis was shown Table S1 in the Supplement 3.

Of the 815 patients included, 396 (48.6%) had at least one CMB, with 210 in the early treatment group and 186 in the delayed treatment group. The remaining 419 patients had no CMBs, distributed nearly equally between the early (210) and delayed (209) treatment groups. Among patients without CMBs, those in the early treatment group were slightly older. Additionally, patients with CMBs in the delayed treatment group were more likely to exhibit small artery disease. Apart from these distinctions, baseline characteristics were comparable between the two treatment groups for both patients with and without CMBs (Table 1).

**Table 1.** Baseline Characteristics of Enrolled Patients According to Cerebral Microbleed (CMB) Presence.

| Characteristics | Without CMBs (n=419) |  |  | With CMBs (n=396) |  |  |
| --- | --- | --- | --- | --- | --- | --- |
|  | Early treatment (n=210) | Delayed treatment (n=209) | P value | Early treatment (n=210) | Delayed treatment (n=186) | P value |
| Age, mean (SD), y | 62.0 (9.5) | 60.0 (9.6) | 0.03 | 64.0 (10.1) | 63.7 (9.0) | 0.81 |
| Male, n (%) | 126 (60.0) | 137 (65.6) | 0.24 | 145 (69.1) | 127 (68.3) | 0.87 |
| Median BMI (IQR), kg/m<sup>2</sup> | 25.3<br>(23.4-27.5) | 24.9<br>(22.9-27.3) | 0.30 | 24.8<br>(22.8-27.1) | 24.8<br>(22.7-27.1) | 0.92 |
| Blood pressure at entry, mean (SD), mm Hg |  |  |  |  |  |  |
| Systolic | 162.7<br>(14.2) | 163.6<br>(15.0) | 0.50 | 163.0<br>(15.2) | 164.7 (14.4) | 0.25 |
| Diastolic | 90.4 (8.7) | 92.1(9.6) | 0.05 | 91.8 (9.2) | 92.4 (10.0) | 0.54 |
| NIHSS score, median (IQR) | 4 (2-6) | 4 (2-5) | 0.72 | 4 (2-6) | 4 (2-6) | 0.73 |
| Medical history, n (%) |  |  |  |  |  |  |
| Hypertension | 157 (74.8) | 154 (73.7) | 0.80 | 164 (78.1) | 150 (80.7) | 0.53 |
| Diabetes mellitus | 59 (28.1) | 57 (27.3) | 0.85 | 51 (24.3) | 52 (28.0) | 0.41 |
| Dyslipidemia | 7 (3.3) | 5 (2.4) | 0.56 | 8 (3.8) | 7 (3.8) | 0.98 |
| Previous stroke | 36 (17.1) | 34 (16.3) | 0.81 | 66 (31.4) | 50 (26.9) | 0.32 |
| Previous TIA | 2 (1.0) | 2 (1.0) | 1.00 | 0 | 0 | - |
| Coronary heart disease | 14 (6.7) | 13 (6.2) | 0.85 | 17 (8.1) | 13 (7.0) | 0.68 |
| Current smoking, n (%) | 80 (38.1) | 86 (41.2) | 0.52 | 79 (37.6) | 63 (33.9) | 0.44 |
| Current drinking, n (%) | 50 (23.8) | 60 (28.7) | 0.25 | 54 (25.7) | 49 (26.3) | 0.89 |
| Previous medications use, n (%) |  |  |  |  |  |  |
| Antihypertensive therapy | 117 (55.7) | 107 (51.2) | 0.35 | 117 (55.7) | 115 (61.8) | 0.22 |
| Antidiabetic therapy | 55 (26.2) | 45 (21.5) | 0.26 | 45 (21.4) | 45 (24.2) | 0.51 |
| Lipid-lowering therapy | 11 (5.2) | 14 (6.7) | 0.53 | 25 (11.9) | 14 (7.5) | 0.14 |
| Antiplatelet therapy | 17 (8.1) | 19 (9.1) | 0.72 | 29 (13.8) | 26 (14.0) | 0.96 |
| Time from onset to randomization, median (IQR), hours | 37.1<br>(29.0-45.0) | 38.0<br>(29.7-45.2) | 0.77 | 38.2<br>(31.0-45.0) | 36.5<br>(28.7-44.0) | 0.07 |
| Time from onset to intervention, median (IQR), days | 1.5<br>(1.2-1.9) | 8.6<br>(8.2-8.9) | <0.001 | 1.6<br>(1.3-1.9) | 8.5 (8.2-8.8) | <0.001 |
| Time from onset to imaging, median (IQR), hours | 33.5<br>(22.8-51.0) | 35.0<br>(24.0-59.0) | 0.10 | 36.4(21.0-75.0) | 34.0<br>(21.0-55.0) | 0.29 |
| Time from randomization to imaging, median (IQR), hours | -2.0(-18.0-20.3) | -1.7(-16.2-24.5) | 0.15 | -2.0(-18.7-42.0) | -1.0(-16.0-18.9) | 0.77 |
| Etiology of stroke, n (%) |  |  | 0.59 |  |  | 0.01 |
| Large-artery atherosclerosis | 54 (25.7) | 59 (28.2) |  | 77 (36.7) | 43 (23.1) |  |
| Small-artery disease | 149 (71.0) | 146 (69.9) |  | 127 (60.5) | 135 (72.6) |  |
| Others | 7 (3.3) | 4 (1.9) |  | 6 (2.9) | 8 (4.3) |  |
| Imaging features, n (%) |  |  |  |  |  |  |
| Moderate-severe WMH* | 86 (41.0) | 82 (39.6) | 0.78 | 151 (72.6) | 131 (70.8) | 0.69 |
| Lacune | 91 (43.3) | 95 (45.9) | 0.60 | 157 (75.9) | 142 (77.2) | 0.76 |
| Moderate-severe EPVS† | 74 (35.4) | 78 (38.2) | 0.55 | 113 (56.2) | 87 (48.1) | 0.11 |
| ICAS | 98 (46.7) | 96 (45.9) | 0.88 | 105 (50.0) | 79 (42.5) | 0.13 |

Abbreviations: CMBs, cerebral microbleed; SD, standard deviation; BMI, body mass index (calculated as weight in kilograms divided by height in meters squared); IQR, interquartile range; NIHSS, National Institute of Health Stroke Scale; TIA, transient ischemic attack; WMH, white matter hyperintensity; EPVS, enlarged perivascular spaces; ICAS, intracranial artery stenosis.
\*Defined as a total Fazekas score between 3 and 6.
†Defined as >10 in either basal ganglia or centrum semiovale.

Patients with CMBs were, on average, older (median age 63.9 vs. 61 years, *P*<0.001) and had a higher prevalence of previous stroke as well as prior use of lipid-lowering or antiplatelet therapy compared to those without CMBs. Furthermore, patients with CMBs more frequently exhibited severe WMHs, lacunes, and EPVSs on MRI (Table S2 in Supplement 3).

Among patients with CMBs, disease severity was mild in 207 patients (52.3%) and moderate-severe in 188 (47.5%). The location of CMBs was strictly deep in 191 patients (48.2%), strictly lobar in 65 (16.4%), and mixed in 140 (35.4%). Patients with moderate-severe CMB burden had a higher rate of hypertension, prior stroke, and previous use of antihypertensive, lipid-lowering, and antiplatelet therapies. These patients also exhibited more advanced CSVD, characterized by severe WMHs, lacunes, and EPVSs, with similar features observed in those with mixed-location CMB. Patients with strictly lobar CMBs were predominantly male, more frequently categorized as having large artery atherosclerotic stroke, and exhibited a longer time from stroke onset to imaging (Table S2 and S3 in Supplement 3).

### Blood pressure reduction

Within 24 hours after randomization, mean (SD) SBP was decreased by 15.7 (18.6) mm Hg (9.2%) and 15.7 (16.2) mm Hg (9.3%) in the early treatment group and by 7.8 (16.7) mm Hg (4.5%) and 7.4 (15.8) mm Hg (4.2%) in the delayed group for patients with and without CMBs (*p* < 0.001 for group difference), respectively. At day 7, the SBP differences between the 2 treatment groups were -11.8 (95% CI, -15.12 to -8.48) in patients with CMBs and -13.64 (95% CI, -16.78 to -10.49) mm Hg in those without CMBs. After starting antihypertensive medications on day 8 in the delayed treatment group, the mean SBP difference between the two groups significantly narrowed to -0.82 and -2.70 mm Hg at day 21 and to -0.82 and -2.14 mm Hg at day 90 for patients with and without CMBs, respectively (Figure 1; Table S4 in Supplement 3).

**Figure 1.**
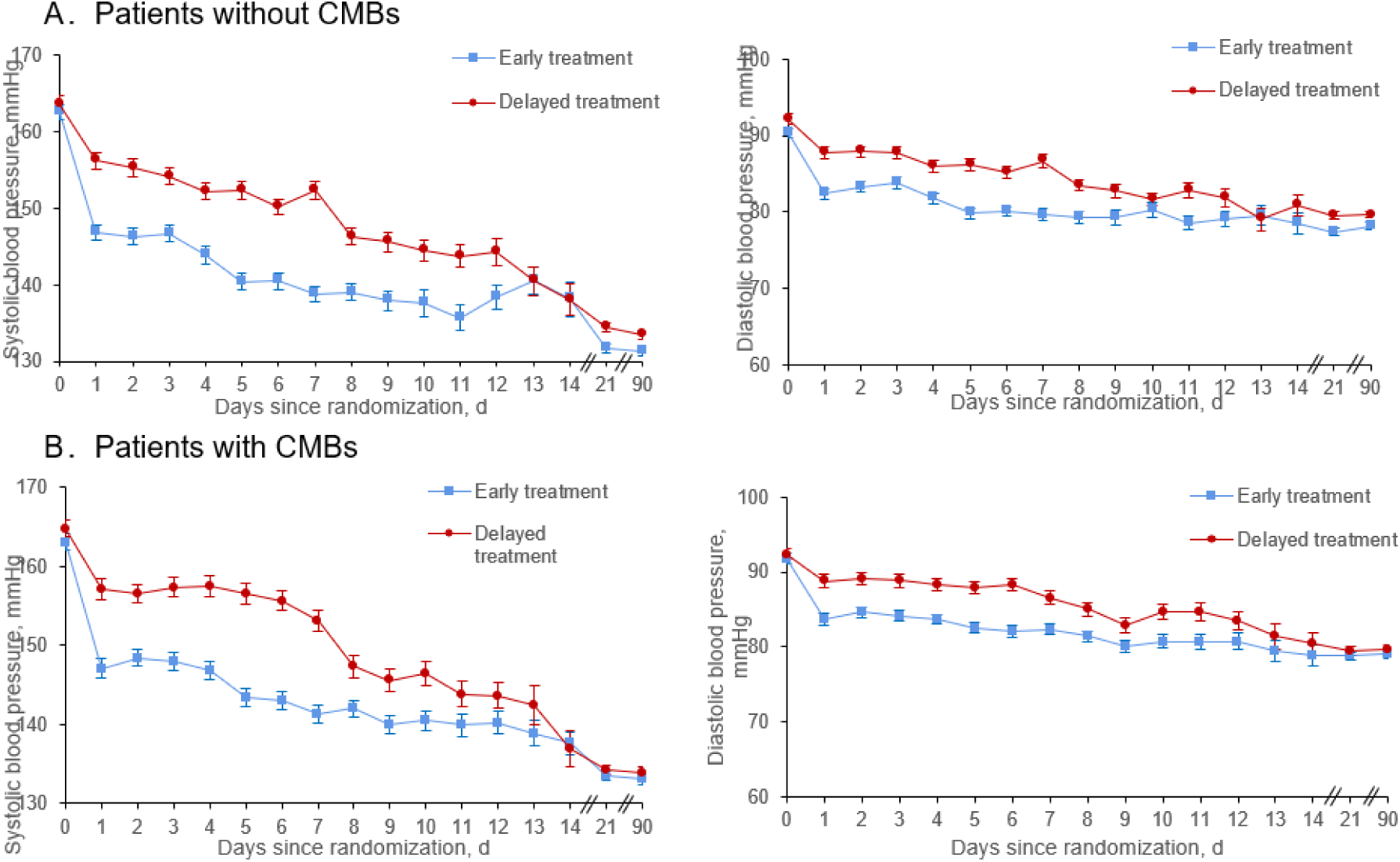
Mean Systolic and Diastolic Blood Pressure (BP) Since Randomization by Status of Cerebral Microbleeds (CMBs) Abbreviations: CMBs, cerebral microbleeds. Three blood pressure measurements were obtained every 3 hours for the first 24 hours and then every 8 hours during hospital admission until day 14 or hospital discharge. In addition, 3 blood pressure measurements were obtained at 21-day and 90-day follow-up visits in all patients. Error bars represent 95% CIs.

### Associations with clinical outcomes

At the 90-day follow-up, 85 of 814 patients (10.4%) experienced the primary outcome of functional dependency or death. Patients with CMBs had a significantly higher risk of functional dependency or death compared to those without CMBs (13.2% vs 7.9%; OR, 1.77; 95%CI, 1.12-2.81; *P*=0.01). Among patients with strictly deep CMBs, the proportion with functional dependency or death was even higher compared to those without CMBs (15.3% vs 7.9%; OR, 2.11; 95%CI, 1.24-3.59; *P*=0.01). This association between strictly deep CMBs and the primary outcome remained significant in adjusted analyses (aOR, 1.92; 95%CI, 1.08-3.40; *P*=0.03). No significant associations were observed between the presence of CMBs and secondary outcomes, including ordinal mRS scores, recurrent stroke and major vascular events (Table 2).

**Table 2.**
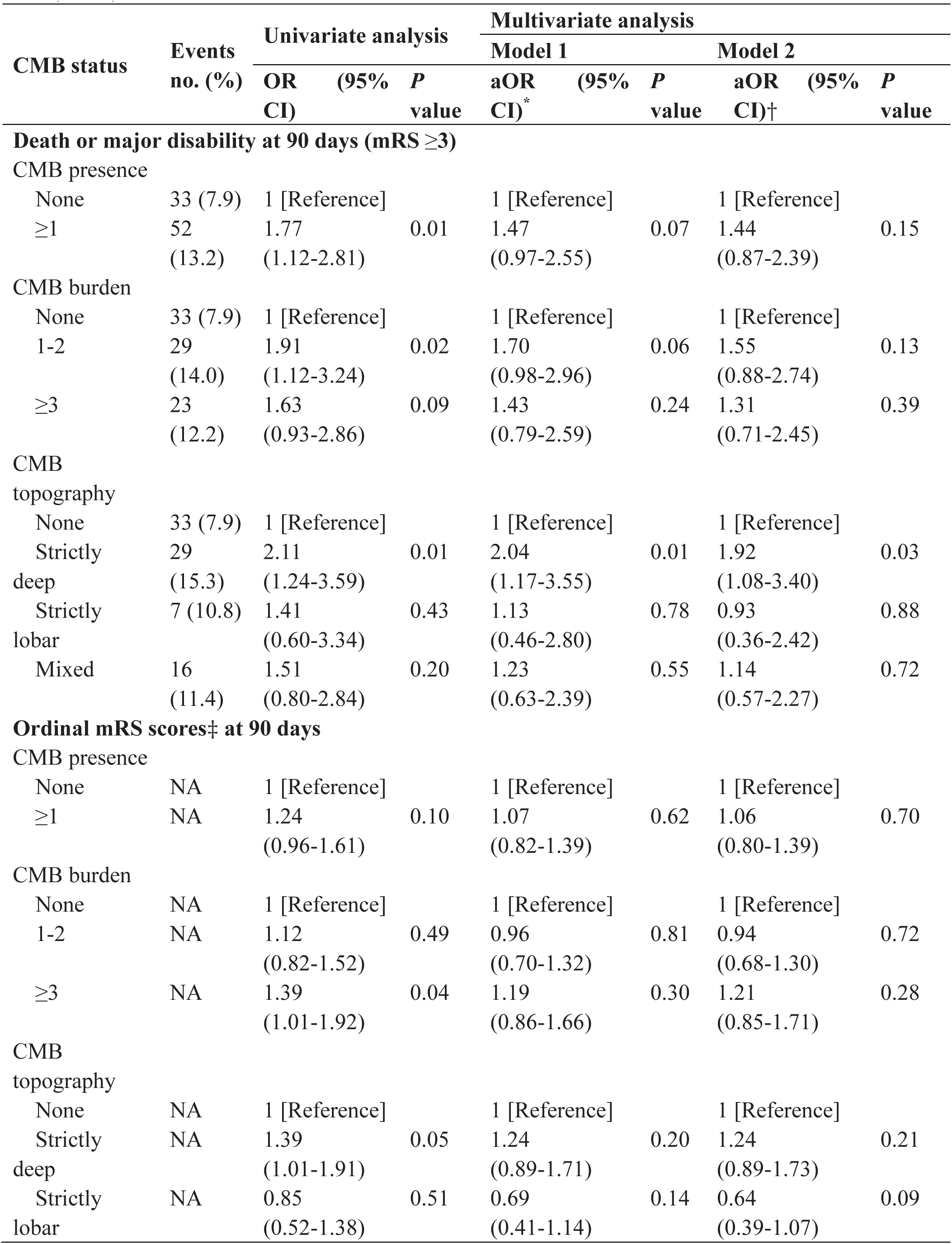
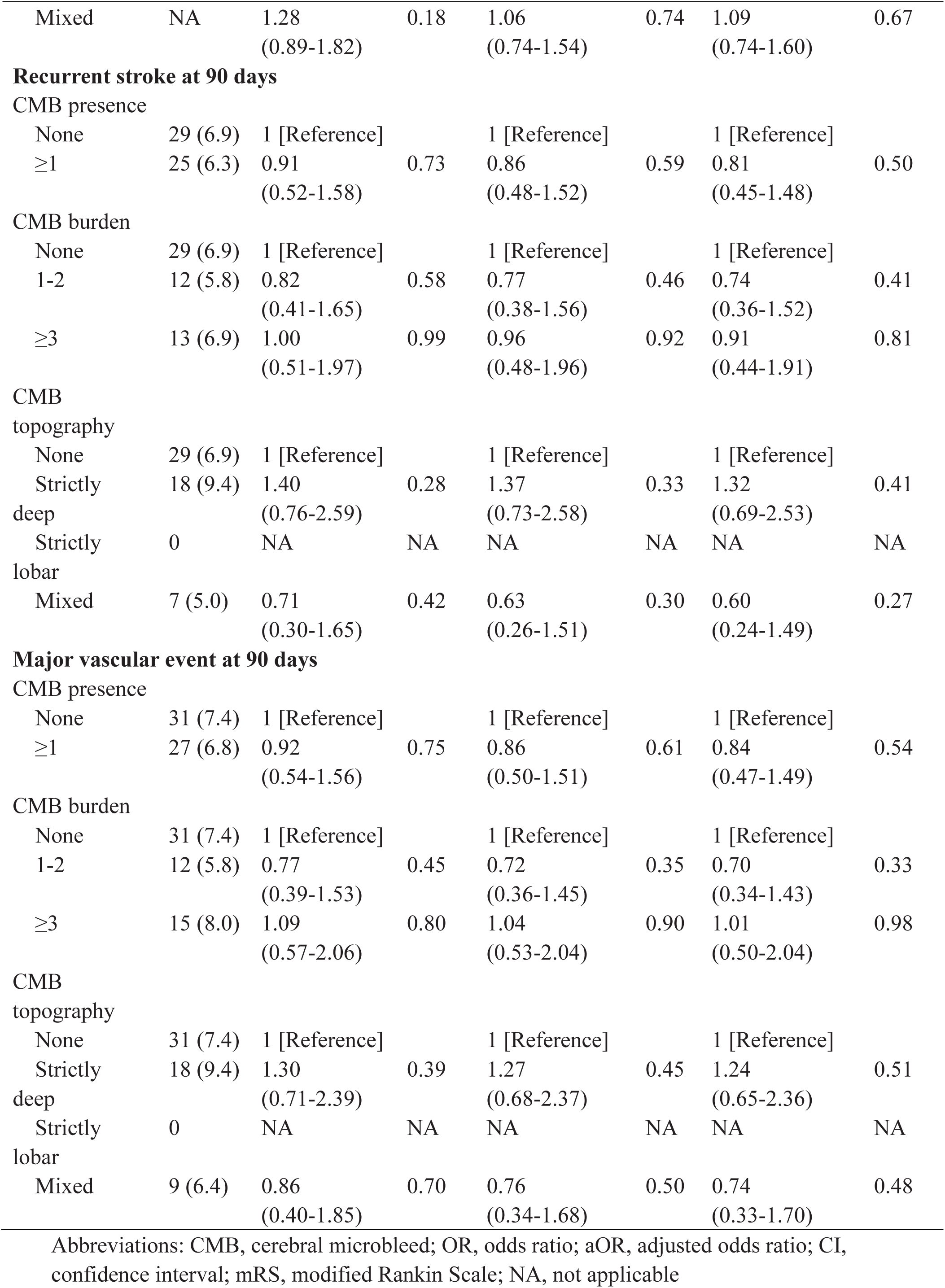
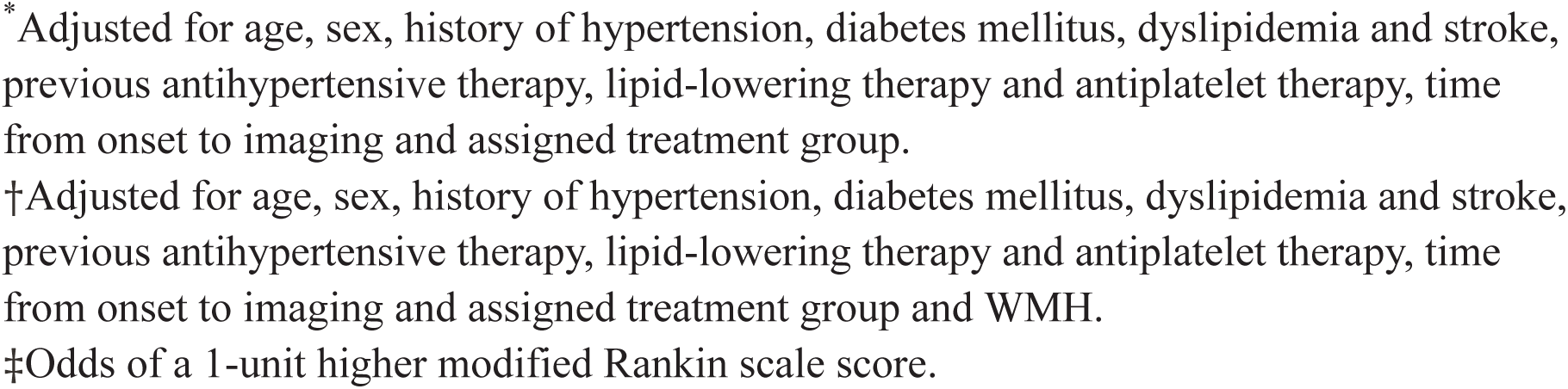
Logistic Regression Analyses of Outcomes According to Cerebral Microbleed (CMB) Status.

### Effect of antihypertensive treatment

Among patients with CMBs, those in the early treatment group had significantly higher odds of 90-day death or functional dependency compared with the delayed treatment group (17.1% vs 8.7%; aOR, 2.17; 95%CI, 1.16-4.07; *P*=0.02). However, this significant increase was not observed in patients without CMBs (8.1% vs 7.7%; aOR, 0.90; 95%CI, 0.43-1.88; *P*=0.79). Patients with moderate-severe CMB burden and those with strictly deep CMBs in the early treatment group also demonstrated higher rates of the primary outcome compared to the delayed treatment group (17.5% vs 6.6%; aOR, 2.85; 95%CI, 1.06-7.67; *P*=0.04 and 20.8% vs 9.0%; aOR, 2.76; 95%CI, 1.15-6.64; *P*=0.02, respectively). Nonetheless, no significant interactions were observed between CMB presence, burden, and topography and antihypertensive treatment (all *P*_interaction_ > 0.05). Similarly, no significant heterogeneity in the treatment effect was identified for secondary outcomes across different CMB subgroups (all *P*_interaction_ > 0.05) (Figure 2).

**Figure 2.**
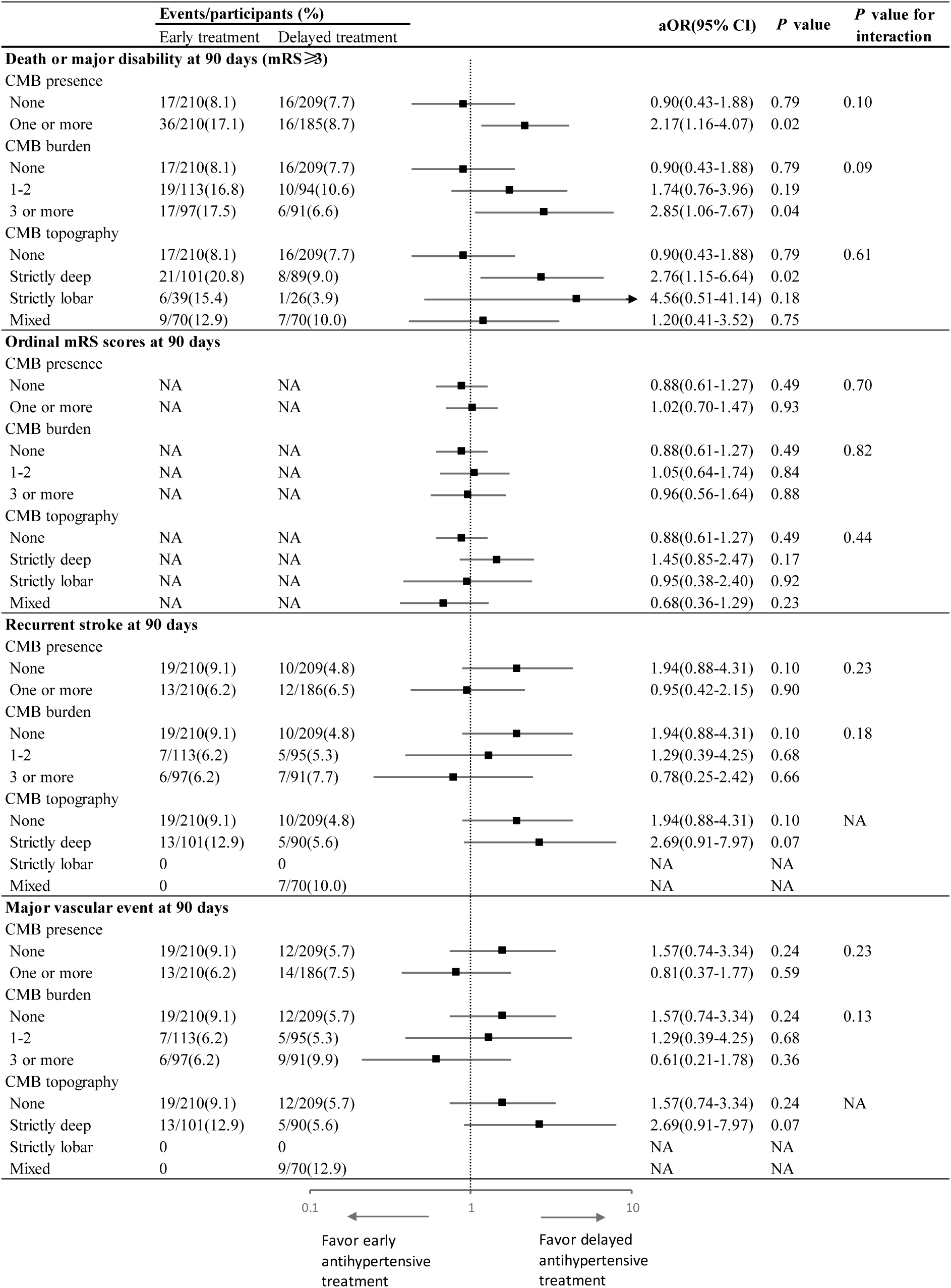
Effects of Early Treatment vs Delay Treatment on Outcomes According to Cerebral Microbleed (CMB) Status. Abbreviations: CMB, cerebral microbleed; aOR, adjusted odds ratio; CI, confidence interval; mRS, modified Rankin Scale; NA, not applicable. Odds ratios were adjusted for age, sex. Odds ratios for ordinal mRS scores represented odds of a 1-unit higher mRS score.

## Discussion

In this subgroup analysis of the CATIS-2 trial, we found a high prevalence of CMBs among AIS patients. The presence of strictly deep CMBs emerged as an independent predictor of 90-day mortality and functional dependency. Importantly, early antihypertensive therapy was associated with a statistically significant increase in the risk of death or functional dependency in patients with CMBs, particularly in those with moderate-to-severe CMB burden and strictly deep CMBs. Despite these findings, no statistically significant interactions were observed between early antihypertensive therapy and CMB status, suggesting the need for further investigation to understand the implications of CMB burden and location on treatment outcomes.

In our study, CMBs were present in 48% of ischemic stroke patients, a prevalence potentially higher than rates observed in other group, as highlighted in a meta-analysis by Wilson et al.^9^ Our cohort also exhibited a higher proportion of strictly deep CMBs (48.2%) and a lower percentage of strictly lobar CMBs (16.4%). Consistent with recent meta-analysis,^22^ Eastern populations had higher odds of deep or mixed CMBs compared to other populations. This disparity may reflect differences in underlying risk factor profiles, including a higher prevalence of hypertension in our study.

In line with our hypothesis, the presence of CMBs in AIS patients was associated with a numerical increase in 90-day mortality or functional dependency. However, evidence regarding the impact of CMBs on functional outcomes in AIS patients remains conflicting.^8,12,13,23,24^ For instance, an exploratory analysis of the New Approach Rivaroxaban Inhibition of Factor Xa in a Global Trial Versus ASA to Prevent Embolism in Embolic Stroke of Undetermined Source (NAVIGATE ESUS) trial suggested that the presence of CMBs in patients with ischemic stroke may indicate generally worse clinical outcomes. Conversely, a subgroup analysis of the Efficacy and Safety of MRI-Based Thrombolysis in Wake-Up Stroke (WAKE-UP) trial found no significant impact of CMBs on 90-day functional outcomes.^23^ Similarly, the Secondary Prevention of Small Subcortical Strokes (SPS3) trial did not identify CMBs as a predictor of mortality in patients with lacunar stroke.^20^ These discrepancies may arise from differences in stroke characteristics, subtype and interventions. Moreover, data from SPS3 trial indicated that CMBs significantly increased the risk of stroke recurrence in patients with lacunar stroke. Several meta-analyses have also reported an elevated risk of stroke recurrence in AIS patients with CMBs.^25,26^ Of note, our study did not identify a significant association between CMBs and stroke recurrence, suggesting that a longer follow-up period may be required to fully elucidate the role of CMBs in stroke recurrence.

Intriguingly, our study found that early antihypertensive therapy was associated with a higher risk of 90-day mortality or functional dependency in AIS patients with CMBs, with a more pronounced effect observed in those with a higher CMB burden and strictly deep CMBs. Data from the SPS3 trial suggest that patients with CMBs may benefit from long-term intensive BP reduction targets.^20^ However, our findings indicate that initiating BP-lowering treatment in the acute phase might have adverse effects in AIS patients with CMBs. One plausible explanation is that cerebral autoregulation (CA) may already be impaired in these, patients, particularly those with multiple CMBs.^15,16^ This impairment could exacerbate CA dysfunction following AIS,^27–30^ leading to tissue hypoperfusion, cerebral ischemia, and subsequently poor functional outcomes. Alternatively, patients with CMBs might represent a subgroup with long-standing poor BP control, characterized by severe arteriosclerosis and reduced adaptability to BP fluctuations. This could account for the worse prognosis observed in patients with deep CMBs—commonly linked to hypertensive vascular disease—following early antihypertensive intervention.

The key contribution of our study is elucidating the efficacy of early versus delayed antihypertensive treatment in AIS patients, with stratification based on baseline CMB status, using data from a high-quality randomized controlled trial. Although no statistically significant treatment interactions were found between treatment assignment and CMBs, a trend was observed suggesting that patients with CMBs, particularly those with higher burden of CMBs and deep CMBs, were associated with higher risk of unfavorable neurological outcome. These findings highlight the need for further investigation in larger cohorts to better understand the hemodynamic mechanisms underlying these observations. Our results suggest potential therapeutic implications that early antihypertensive treatment should be approached with caution in AIS patients with CMBs. This insight underscores the importance of individualized BP management strategies during the acute phase of ischemic stroke, tailored to specific imaging characteristics and patient profiles.

This study has several limitations. First, our results were constrained by the limited availability of MRI sequences, with only approximately 17% of CATIS-2 trial participants ultimately included, which limits the generalizability of our findings. Second, the non-standardization of GRE or SWI sequence acquisition parameters, along with the absence of MRI parameter data, may have caused heterogeneity in CMB detection rates across recruitment sites and potentially confounded our findings. This further highlights the need for harmonized imaging protocols across study sites in future trials. Third, the relatively low rate of observed primary outcomes and the small number of events for bleeding complications limited our statistical power and our ability to study the effect of CMBs on bleeding risk. Fourth, our study predominantly included patients with minor strokes and excluded those who received intravenous thrombolysis or endovascular thrombectomy at baseline. Therefore, our findings may not be generalizable to patients receiving reperfusion therapy or those with severe strokes.

## Conclusions

In this subgroup analysis of the CATIS-2 trial, the presence of strictly deep CMBs was independently associated with increased 90-day mortality and functional dependency. Notably, patients with CMBs demonstrated poorer functional outcomes following early antihypertensive therapy, with the effect being more pronounced in those with a higher CMB burden and deep CMBs. Although no significant interaction was identified between CMB status and early antihypertensive treatment, the observed trends highlight the need for caution when initiating early antihypertensive therapy in this population. Further studies are warranted to validate these findings and to optimize BP management strategies for AIS patients with CMBs.

## Acknowledgment

The authors thank the study participants and their relatives and the clinical staff at all participating hospitals for their support and contributions to this project.

## Supplemental Material

Study Protocol Statistical Analysis Plan Supplement 3 CONSORT Checklist

